# ‘Vivaldi’: An amplicon-based whole genome sequencing method for the four seasonal human coronaviruses 229E, NL63, OC43 & HKU1, alongside SARS-CoV-2’

**DOI:** 10.1101/2024.06.26.24308604

**Authors:** C. Patrick McClure, Theocharis Tsoleridis, Nadine Holmes, Joseph G. Chappell, Timothy Byaruhanga, Joshua Duncan, Miruna Tofan, Abdul Khater, Louise Berry, Gemma Clark, William L. Irving, Alexander W. Tarr, Jonathan K. Ball, Stuart Astbury, Matt Loose

## Abstract

Prior to the emergence of SARS-CoV-2 in 2019, Alphacoronaviruses 229E and NL63 and Betacoronaviruses OC43 and HKU1 were already established endemic ‘common cold’ viral infections. Despite their collective contribution towards global respiratory morbidity and mortality and potential to inform the future trajectory of SARS-CoV-2 endemicity, they are infrequently sequenced. We therefore developed a 1200bp amplicon-based whole genome sequencing scheme targeting all four seasonal coronaviruses and SARS-CoV-2.

The ‘Vivaldi’ method was applied retrospectively and prospectively using Oxford Nanopore Technology to approximately 400 seasonal coronavirus infections diagnosed in Nottingham, UK, from February 2016 to July 2023. We demonstrate that the amplicon multiplex strategy can be applied agnostically to determine complete genomes of five different species from two coronaviral genera. 304 unique seasonal coronavirus genomes of greater than 95% coverage were achieved: 64 for 229E, 85 for NL63, 128 for OC43 and 27 for HKU1. They collectively indicated a dynamic seasonal coronavirus genomic landscape, with co-circulation of multiple variants emerging and declining over the UK winter respiratory infection season, with further geographical distinction when compared to a global dataset. Prolonged infection with concomitant intra-host evolution was also observed for both Alpha-(NL63) and Betacoronaviruses (OC43).

This data represents the largest single cohort of seasonal coronavirus genomes to date and also a novel amplicon scheme for their future global surveillance suitable for widespread and easy adoption in the post-SARS-CoV-2 era of viral genomics.

## Introduction

The *Coronaviridae* family belongs to the *Nidovirales* order of positive single-stranded RNA viruses with relatively large genomes (27-33 kb) and four genera (*Alpha*-, *Beta*-, *Gamma*- and *Deltacoronavirus*) within the Orthocoronavirinae subfamily, with Alpha- and Betacoronavirus genera members capable of human infection (1, 2).

Prior to the emergence of SARS-CoV-2 in 2019, four other Coronaviruses – Alphacoronaviruses 229E, NL63 and Betacoronaviruses OC43 and HKU1– had previously established endemic status (3–6). Collectively these are responsible for a significant proportion of global respiratory infections as part of the informal ‘common cold’ viral grouping (7). Whilst they present a broad spectrum of disease severity in all age groups, infections are typically but not exclusively symptomatically mild (8), which has in part led to their clinical and genomic under-investigation to the detriment of understanding the emergence and future trajectory of SARS-CoV-2 endemicity (7, 9, 10).

To date, six 229E genotypes (1–6) and three for NL63 (A-C) (11, 12) have been described for the Alphacoronaviruses, whilst Betacoronaviruses OC43 and HKU1 have been categorised into 10 (A-K) and 3 (A to C) genotypes, respectively (12–16). Genetic diversity is also generated by extensive recombination events within and between genera (11, 15, 17).

Amplicon-based Whole Genome Sequencing (WGS), utilising contiguous overlapping PCR products of various size, has previously been described for a wide range of viruses (18–24), yielding both population-level epidemiological insight and outbreak management with near-realtime capability (19). There is a trade off in the priming strategy between the increased sensitivity of smaller amplicon tiling versus the heightened chance of primer mismatch where greater number of primers are used. Greater diversity in the targeted viral taxon, as observed in established endemic pathogens, creates a more challenging target preferentially necessitating fewer amplicons to span the genome (18, 24). Continuous viral evolution, especially noticeable during the early phases of rapid viral spread following a recent zoonotic spillover can result in primer mismatches, necessitating careful surveillance for amplicon ‘drop-out’ and swift primer redesign (25).

Due to the perceived low medical threat posed by seasonal coronaviruses their inclusion in diagnostic panels and molecular epidemiological surveillance programmes has been overlooked in some instances. Since the emergence of SARS-CoV-2, there is increasing interest in the seasonal coronavirus molecular and clinical epidemiology (10, 26–28).

This study describes an amplicon-based approach to provide whole genome sequences of the four seasonal coronaviruses alongside SARS-CoV-2. We have applied the methodology both retrospectively to a large cohort of archival extracts pre-SARS-CoV-2 emergence and prospectively to contemporary post-pandemic infections. The resulting sequence data provides greater depth of insight into seasonal coronaviral variation, potentially informing future SARS-CoV-2 endemicity. Contemporary viral genomes and future uptake of the methodology will facilitate further understanding of seasonal coronavirus evolution in a post-pandemic immune landscape.

## Methods

### Samples

Surplus total nucleic acid from anonymised Coronaviral-positive patient respiratory samples diagnosed as part of the routine care pathway at Nottingham University Hospitals NHS Trust (NUH NHST) was stored at −70°C from February 2016 to December 2018 and May 2021 to July 2023 as previously described (29, 30). Prior to May 2021, the AusDiagnostics Respiratory 16-plex clinical diagnostic panel used could discriminate seasonal coronavirus type, but this was not always recorded by laboratory personnel, nor was the absolute quantitative template copy number. Due to potential clinical confusion and focussing of diagnostic resources, seasonal coronaviruses were not routinely investigated at the onset of the SARS-CoV-2 Pandemic between February 2020 and April 2021, where after no diagnostic distinction was made between 229E, NL63, OC43 & HKU1 with a newly configured AusDiagnostics respiratory multiplex RT-qPCR in use. Available nucleic acid with a recorded seasonal coronavirus type and a diagnostic laboratory quantitation of greater than 10 copies per 10ul RNA were selected from the 2016 to 2018 archive, with the exception of HKU1, where all positives were tested. Subsequently, all available diagnosed seasonal coronavirus positive extracts were investigated from the 2021 to 2023 sub-cohort. Anonymised diagnostic laboratory PCR results were curated and analysed in Excel. NUH NHST approved extended molecular investigation of diagnosed coronavirus positives under clinical audit number 23-078C.

### Amplicon scheme primer design

All seasonal coronavirus genomes deposited in GenBank by July 2021 were downloaded, aligned as below, and used to construct maximum likelihood trees, from which 5 distinct lineage sequences were chosen and concatenated with different SARS-CoV-2 variant sequences to generate 5 templates for PrimalScheme (21). PrimalScheme was then instructed to generate primer sequences targeting 1200bp regions of the concatenated template, covering all 4 seasonal coronavirus and SARS-CoV-2 genomes. The outputted 278 primer scheme was manually inspected in MEGA7 for mismatch against the alignment of all available genomes and accordingly edited with degeneracy or redesigned where 3’ mismatch was observed in a significant proportion of the global dataset. Amplicon coverage was subsequently reviewed with each sequencing run, redesigning primers where amplicon drop-out or excessively lower coverage was observed. Amplicon balancing was further attempted in a minority of instances by doubling or halving primer concentration where coverage remained significantly low or high respectively. Final selected primer sequences and relative concentrations are listed in supplementary table 1.

### cDNA synthesis, PCR & whole genome sequencing

Coronaviral cDNA was prepared with RNA to cDNA EcoDry™ Premix containing random hexamers (Takara Bio) as per the manufacturer’s instruction, then processed similarly to previously described (20). Briefly, up to 2.5 µl of cDNA was used as template in a 25 µl PCR reaction assembled with Q5® High-Fidelity 2X Master Mix (New England Biolabs) and either primer set 1 (odd numbered primer pairs) or 2 (even numbered primer pairs) of either targeting individual coronaviral species or all 5 combined at a final concentration of 0.015 µM unless stated otherwise (Supplementary table 1). PCR reactions were thermocycled as follows: 98°C for 30 seconds then 45 cycles of 98°C for 15 seconds and 65°C for 5 minutes. PCR products from reaction 1 and 2 were inspected for specificity and yield on a 2% agarose gel with ethidium bromide, and subsequently combined before Qubit quantification and normalisation to 100ng of DNA in 7.5 µl water. Amplicons were prepared and barcoded using the SQK-LSK109 and EXP-NBD196 kits respectively (Oxford Nanopore), and sequenced on an Oxford Nanopore GridION as previously described (19).

### Sequence and phylogenetic analysis

Following basecalling using Guppy (v6.5.7), demultiplexed reads passing the quality threshold (average score >7) were used as the input for the ARTIC pipeline (21). Briefly, reads were filtered to remove those below 700bp and above 1400bp, and aligned to a reference.fasta containing the 4 seasonal coronaviruses and SARS-CoV-2 using Minimap2 (31). A .bed file corresponding to the primers used in the scheme was then used to softmask resulting alignments via the ARTIC align_trim script to ensure variants were not called in primer sites. Finally, the softmasked alignment was used as the input for nanopolish (32) to generate a consensus sequence which was then aligned to the reference using MUSCLE (33). Coverage was based on a 20x individual amplicon threshold with those dropping below this classified as dropouts. The reference .fasta and scheme .bed file are available on request in advance of submission to GenBank.

Genomes were aligned for the presented analysis using the Geneious Prime 2019.0.4 software with the relevant seasonal coronavirus species genomes deposited in GenBank by December 2023. Maximum-Likelihood trees were generated to visualise the evolutionary relationships between high quality (>95% complete) study and publicly available genomes with IQ-TREE2 using the following models of evolution as suggested by the software’s model finder respectively: 229E - TIM+F+R2; NL63 - TIM+F+R3; OC43 - TIM+F+R6; HKU1 - TN+F+R2, with 1000 bootstraps of the Shimodaira–Hasegawa approximate likelihood ratio test (SH-aLRT). Seasonality was assigned for all sequences with >95% coverage, using the first instance of infection in serially sampled patients only. Core season was defined as October to May (e.g. 16/17 for samples sequenced between October 2016 and May 2017), whilst June to September was considered out of typical season (e.g. 2017 for a sequence generated in June 2017). Snip-it plots from the CIVET tool (https://github.com/artic-network/civet) were used to illustrate genetic difference between sequences derived from prolonged infection (34).

### Figure visualisation and data availability

Figures were generated in R v4.3.2 using the package ggplot2 v3.5.1. At time of preprint, sequences are in preparation for submission to GenBank and scheme files are available on request.

## Results

### Seasonal coronavirus diagnosis in a regional UK diagnostic lab

Seasonal coronaviruses represent a significant proportion of the viral respiratory pathogens identified in the Nottingham University Hospitals NHS Trust diagnostic laboratory (NUH NHST). Since their inclusion in the respiratory diagnostic RT-qPCR multiplex in February 2016, through to the study end in July 2023, 1540 positives have been recorded in c. 62,000 tests (data not shown), representing 2.5% of all samples tested and 5.75% of those with a diagnosed viral infection.

Pre-Pandemic, broadly similar numbers of coronaviral positives were identified in each yearly infection season, most abundantly in one of the first 3 months of the year (Figure 1), with a peak seen in March of 2016, February of 2017 and January of both 2018 and 2019. However, infections were recorded all year round, but are significantly decreased to a background level outside of the wider coronaviral season of October to May. Post-pandemic, considerably less samples were submitted for seasonal coronavirus testing due to reconfigured diagnostic practice, reducing discrimination of seasonality. Nonetheless, 2023 appeared to indicate seasonal corona infections peaking in January as previously.

**Figure 1:**
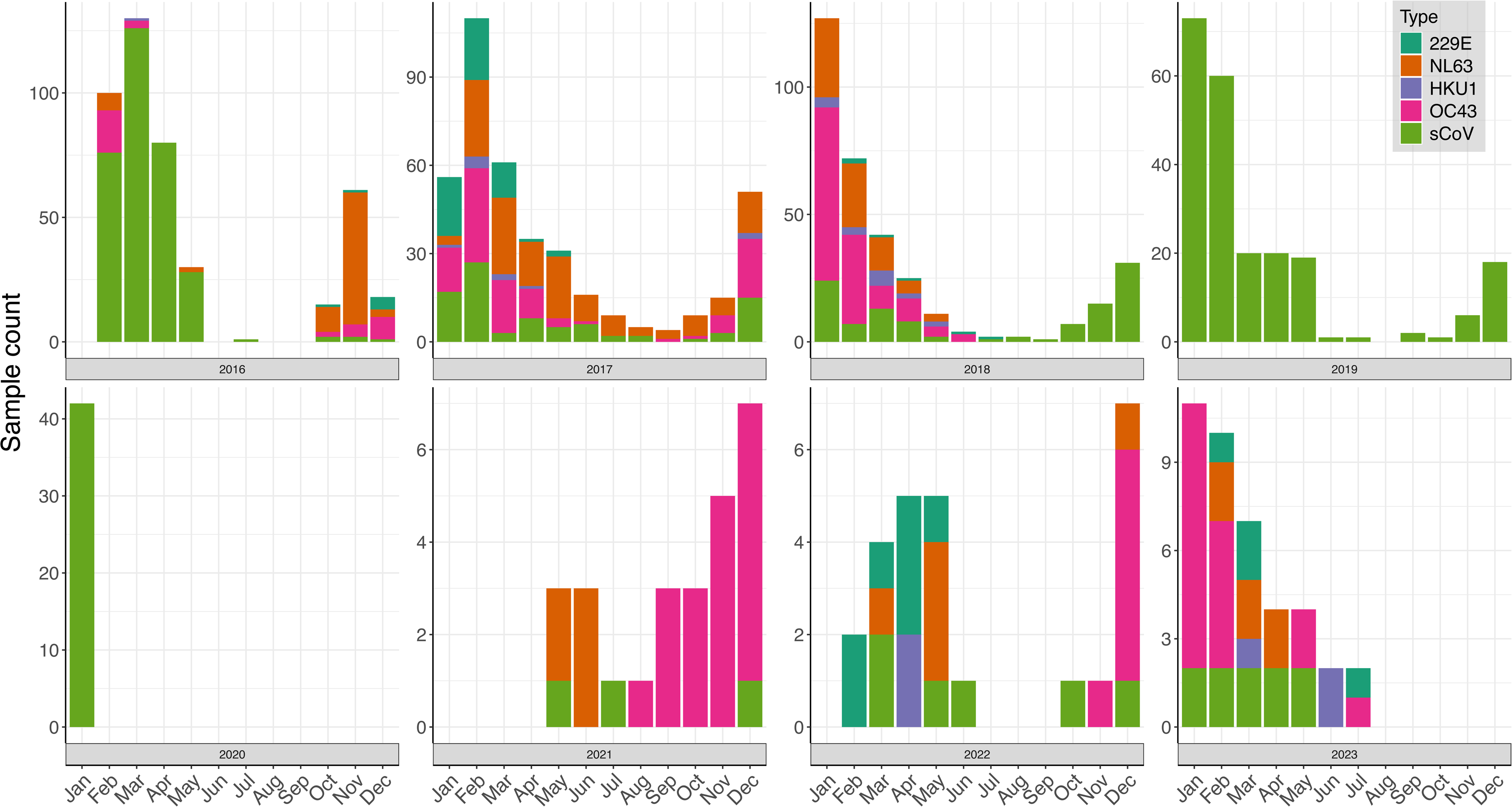
Diagnosed Seasonal Coronavirus infections, Nottingham University Hospitals NHS Trust UK, February 2016 to July 2023. Seasonal coronavirus testing was undertaken with the AusDiagnostics Respiratory 16-plex assay beginning in February 2016 and able to discriminate seasonal coronavirus type, but this was not always recorded, in which case ‘sCoV’ was indicated. In February 2020 Seasonal Coronavirus detection was switched off to avoid clinical confusion at the onset of the SARS-CoV-2 pandemic. Seasonal Coronavirus testing was resumed in May 2021 in a more selective capacity, where after no diagnostic distinction was made between 229E, NL63, OC43 & HKU1 on a newly reconfigured AusDiagnostics panel. All available samples collected post-SARS-CoV-2 emergence were attempted for WGS and where successful the type is pre-sented.

Whilst seasonal coronavirus type was only noted for approximately half (798) of recorded positives (1531), with further periodic unevenness (e.g. mid July onward no discrimination made in laboratory records), OC43 (312) and NL63 (308 indicated in approximately equal numbers. In contrast 229E was observed much more infrequently with 80 positives and HKU1 the rarest of types with just 33 recorded infections. With typing determined by this sequencing study, all four seasonal coronaviruses were again detected post-pandemic, with OC43 the most abundant and in line with more extensive seasonal coronavirus epidemiological studies reported in the UK (7) and USA (10) over similar time periods.

### A method to sequence Seasonal Coronaviruses

To interrogate the molecular epidemiology of the seasonal coronaviruses detected at NUH NHST, a novel 1200bp amplicon sequencing scheme to generate near-complete genomes was developed and applied to available typed and untyped archived nucleic acid extracts.

In total, 402 unique samples were sequenced across 700 reactions, summarised in Table 1 and Figure 2:

**Figure 2:**
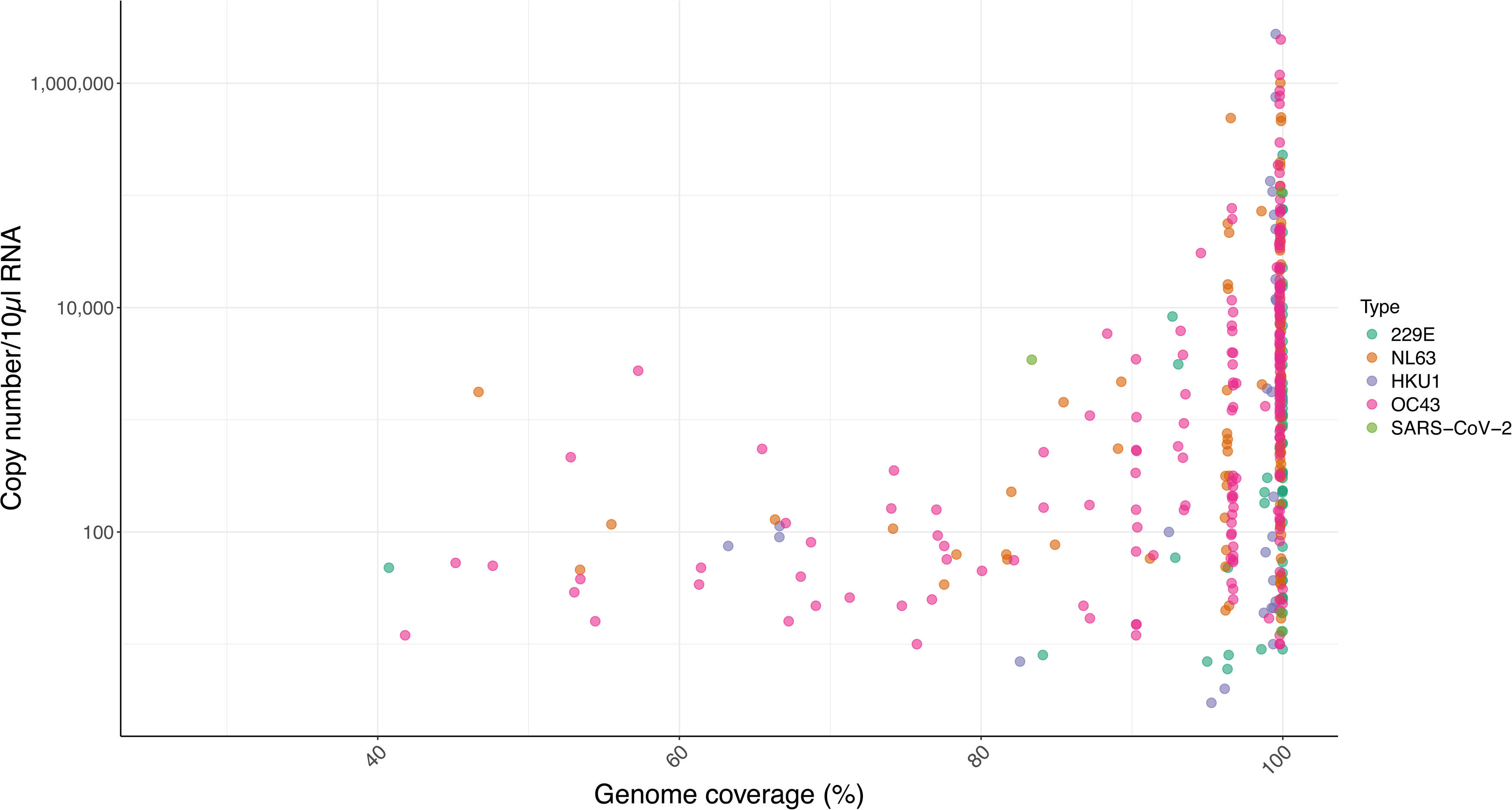
Genome coverage of Seasonal Coronaviruses by the ‘Vivaldi’ 1200bp amplicon scheme. The genome coverage of each Seasonal Coronavirus 229E, NL63, HKU1 & OC43 (x axis) and additionally SARS-CoV-2, expressed as a percentage of the total region targeted by the amplicon scheme is shown compared to the template copy number calculated by the diagnostic assay (Copies per 10 µl RNA, AusDiagnostics, y-axis).

**Table 1:** Summary of Coronaviral sequencing investigations and genomic coverage achieved.

| All sequencing reactions attempted |  |  |  |  |  |  |  |
| --- | --- | --- | --- | --- | --- | --- | --- |
|  |  | Genome coverage |  |  |  |  |  |
| Coronavirus species | total | >90 | % | >95 | % | >99 | % |
| 229E | 115 | 99 | 86.09 | 88 | 76.52 | 72 | 62.61 |
| NL63 | 233 | 178 | 76.39 | 167 | 71.67 | 85 | 36.48 |
| OC43 | 265 | 207 | 78.11 | 166 | 62.64 | 106 | 40.00 |
| HKU1 | 78 | 68 | 87.18 | 54 | 69.23 | 26 | 33.33 |
| SARS-CoV-2 | 9 | 7 | 77.78 | 6 | 66.67 | 5 | 55.56 |
| **Grand total** | **700** | 559 | 81.11 | 481 | 69.35 | 294 | 45.60 |
| Unique samples with final scheme iteration |  |  |  |  |  |  |  |
|  |  | Genome coverage |  |  |  |  |  |
| Coronavirus species | total | >90 | % | >95 | % | >99 | % |
| 229E | 71 | 69 | 97.18 | 64 | 90.14 | 58 | 81.69 |
| NL63 | 100 | 86 | 86.00 | 85 | 85.00 | 65 | 65.00 |
| OC43 | 185 | 149 | 80.54 | 128 | 69.19 | 94 | 50.81 |
| HKU1 | 38 | 29 | 76.32 | 27 | 71.05 | 25 | 65.79 |
| SARS-CoV-2 | 8 | 7 | 87.50 | 6 | 75.00 | 5 | 62.50 |
| **Grand total** | **402** | 340 | 85.51 | 310 | 78.08 | 247 | 65.16 |

The relative higher prevalence of OC43 and NL63 was reflected in both attempted and ultimately unique sequenced genomes. High quality (>95%) genomes were achieved for 69% (OC43) to 90% (229E) of coronaviral samples, whilst complete amplicon coverage (and near complete genomes) were achieved in 51 to 82% of cases. However, all 702 sequenced samples generated at least 25% of the genome, facilitating conclusive typing, although further samples were initially attempted by PCR, but failed to generate any observable amplicons and were thus not progressed to sequencing (data not shown).

Complete genome sequences could be retrieved over the wide dynamic range of recorded diagnostic lab template copy inputs for all 4 seasonal coronavirus types, from as little as 10 copies to in excess of a million (Figure 2, NB not all copy numbers were recorded in the diagnostic lab records). 229E and HKU1 samples with less than 90% coverage had a copy number value of c. 100 copies (per 10µl) template or less. By contrast, NL63 and OC43 samples often yielded sequence data with low coverage, even at relatively high copy number, suggesting primer mismatch as the likely cause Figure 2.

A limited subset of predominantly post-pandemic samples were amplified and sequenced both with all 5 primer sets combined in each pool (The Full Vivaldi, containing 70 and 69 primer pairs in pool 1 and 2 respectively) and in some cases additionally with the conventional single primer sets (Targeted Vivaldi, containing 13 to 15 primer pairs in each pool), (Supplementary Figure and table 1). The Full Vivaldi approach did yield some full genomes but was more prone to produce incomplete genomes compared to the targeted approach. It was, however, still capable of typing seasonal coronavirus positives (Supplementary Figure 1). When tested, all but two of the partial genomes could be converted to near-complete (>96% coverage) by repeating amplification with targeted primers.

A sample positive for both OC43 (3431 copies per 10µl template) and SARS-CoV-2 (959 copies) generated complete coverage for the OC43 genome (UKN23_OC43_5) and a partial SARS-CoV-2 genome (65.95%,) when amplified by the Full Vivaldi method. The SARS-COV-2 genome coverage could be improved to 83.36% (UKN23_CoV2_1) when amplified with the targeted scheme, whilst the OC43 targeted scheme maintained complete coverage.

A small number of patients infected with OC43 were sampled and sequenced within a typical acute phase timeframe of infection and generated paired genomes of >95% coverage, allowing insight into short-term intra-patient virus evolution and accuracy of base-calling across the 30202 sequenced positions. Three paired samples taken one (n=2) and three days apart exhibited no differences, a further two patients saw single differences over two- and seven-day timepoints, whilst the sixth patient presented two mutations in a six-day interval. Taken together, these limited changes are within the expected mutational rate of seasonal coronavirus genomes (35) and indicative of the high fidelity of the amplicon method.

### Genetic epidemiology of seasonal coronaviruses in Nottingham, UK, 2016-2023

The developed amplicon scheme thus generated a considerable number of genomes across the pre- and post-pandemic years at our single regional centre, even when compared to the entire available global dataset. High quality sequences (>95%,) from both the study (64 for 229E, 85 for NL63, 128 for OC43 and 27 for HKU1) and GenBank were aligned and used to construct phylogenetic trees (Figures 3 to 6). Lineages were assigned as per previous studies, summarised in Ye et al (16).

**Figure 3:**
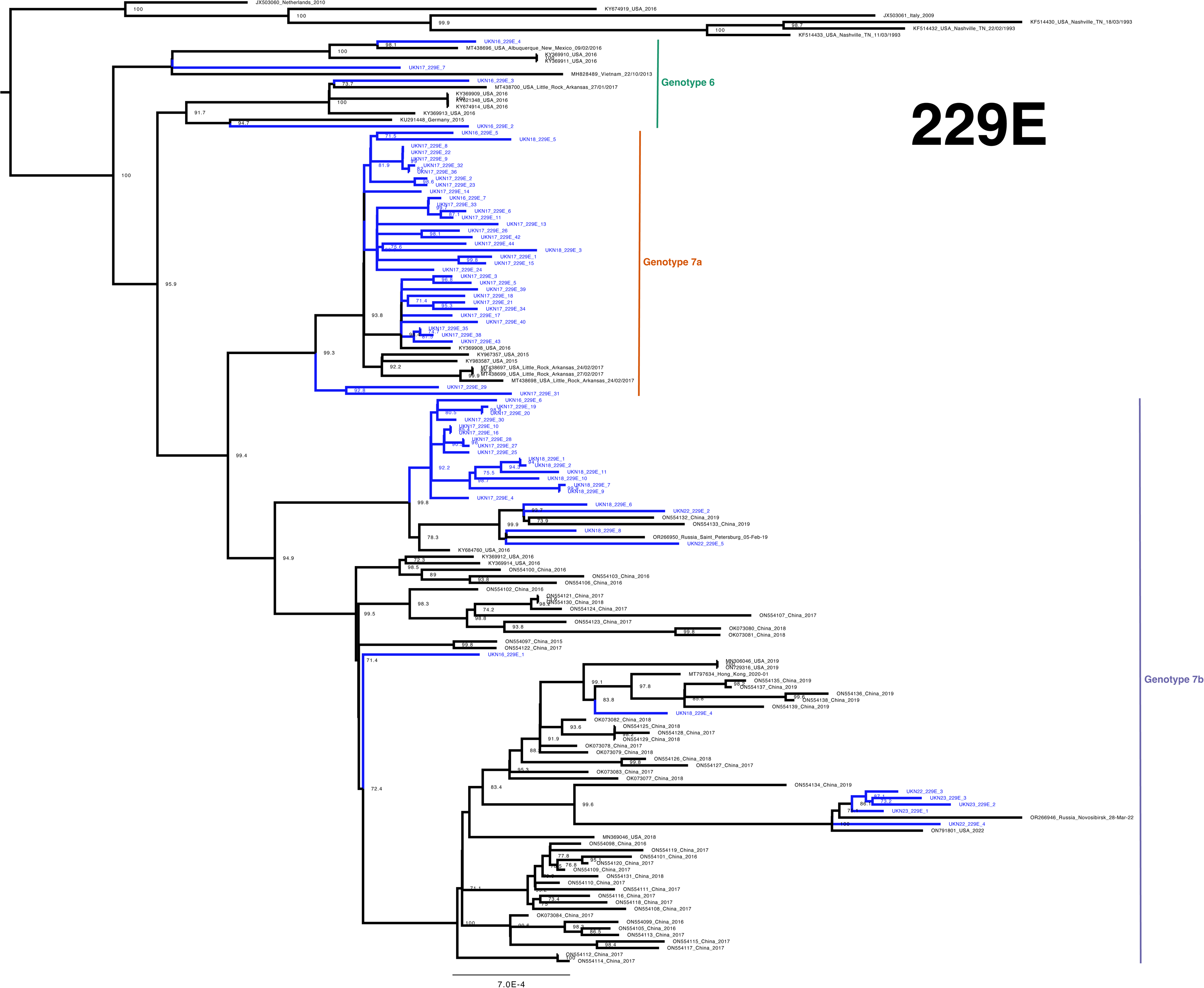
Phylogeny of 229E Seasonal Coronavirus. Phylogenetic relationship by maximum-likelihood method of all Nottingham, UK, study sequences with >95% coverage (blue text) and publicly available genomes (black text) retrieved from GenBank in December 2023 (identified by accession num-ber/country [with region where noted]/year). Numbers to the right of tree nodes indicate SH-aLRT (Shimodaira–Hasegawa approximate likelihood ratio test) branch support, with values <70 not shown. Branch lengths are drawn to a scale of nucleotide substitutions per site, with scale indicated. Tentative geno-types based on well supported clustering (>90 branch support) with lineage exemplar sequences as presented by Ye et al (16) are indicated by vertical bars.

**Figure 4:**
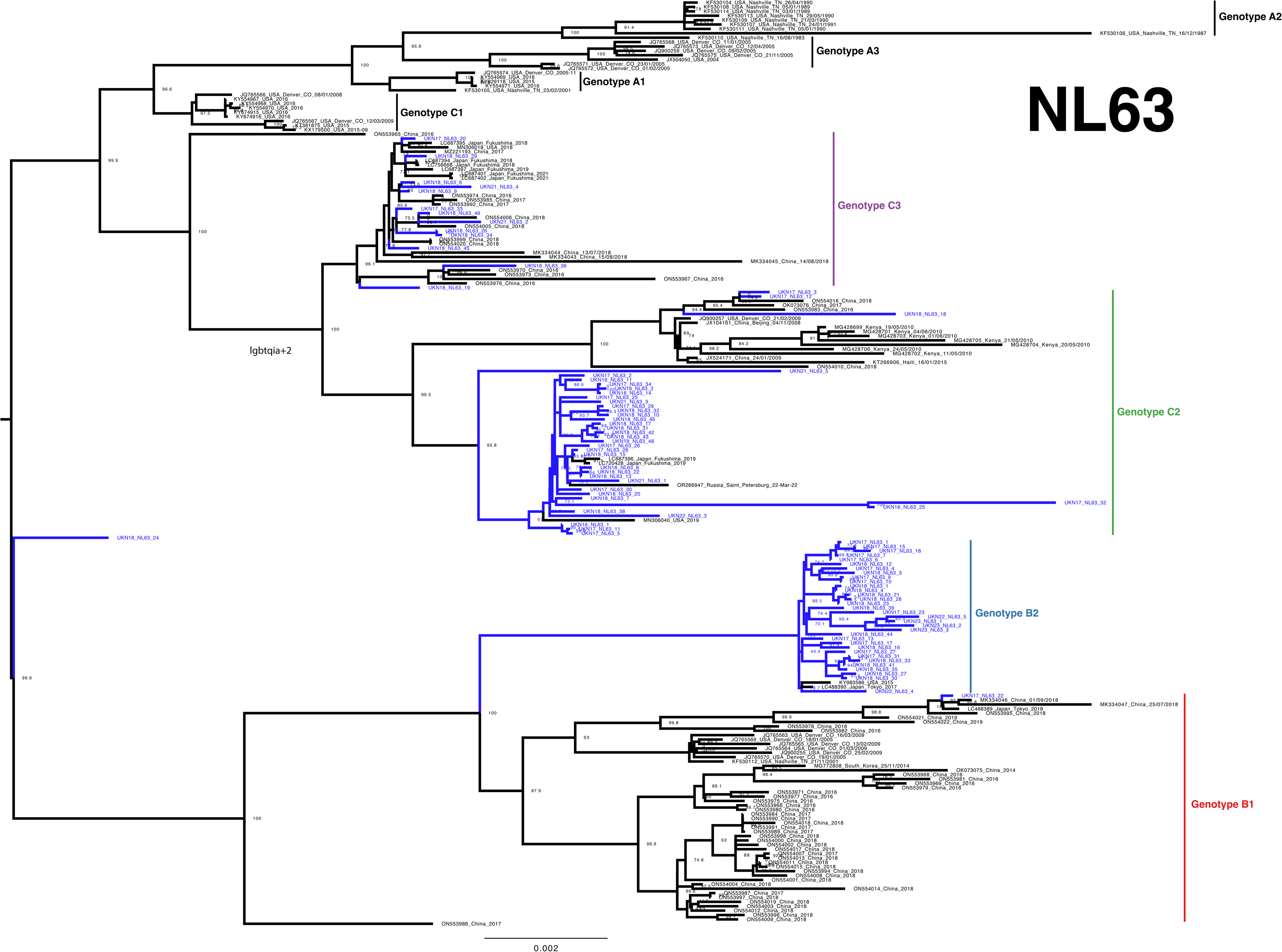
Phylogeny of NL63 Seasonal Coronavirus. Phylogenetic relationship by maximum-likelihood method of all Nottingham, UK, study se-quences with >95% coverage (blue text) and publicly available genomes (black text) retrieved from GenBank in December 2023 (identified by ac-cession number/country [with region where noted]/year). Numbers to the right of tree nodes indicate SH-aLRT (Shimodaira–Hasegawa approximate likelihood ratio test) branch support, with values <70 not shown. Branch lengths are drawn to a scale of nucleotide substitutions per site, with scale indicated. Tentative genotypes based on well supported clustering (>90 branch support) with lineage exemplar sequences as presented by Ye et al (16) are indicated by vertical bars.

**Figure 5:**
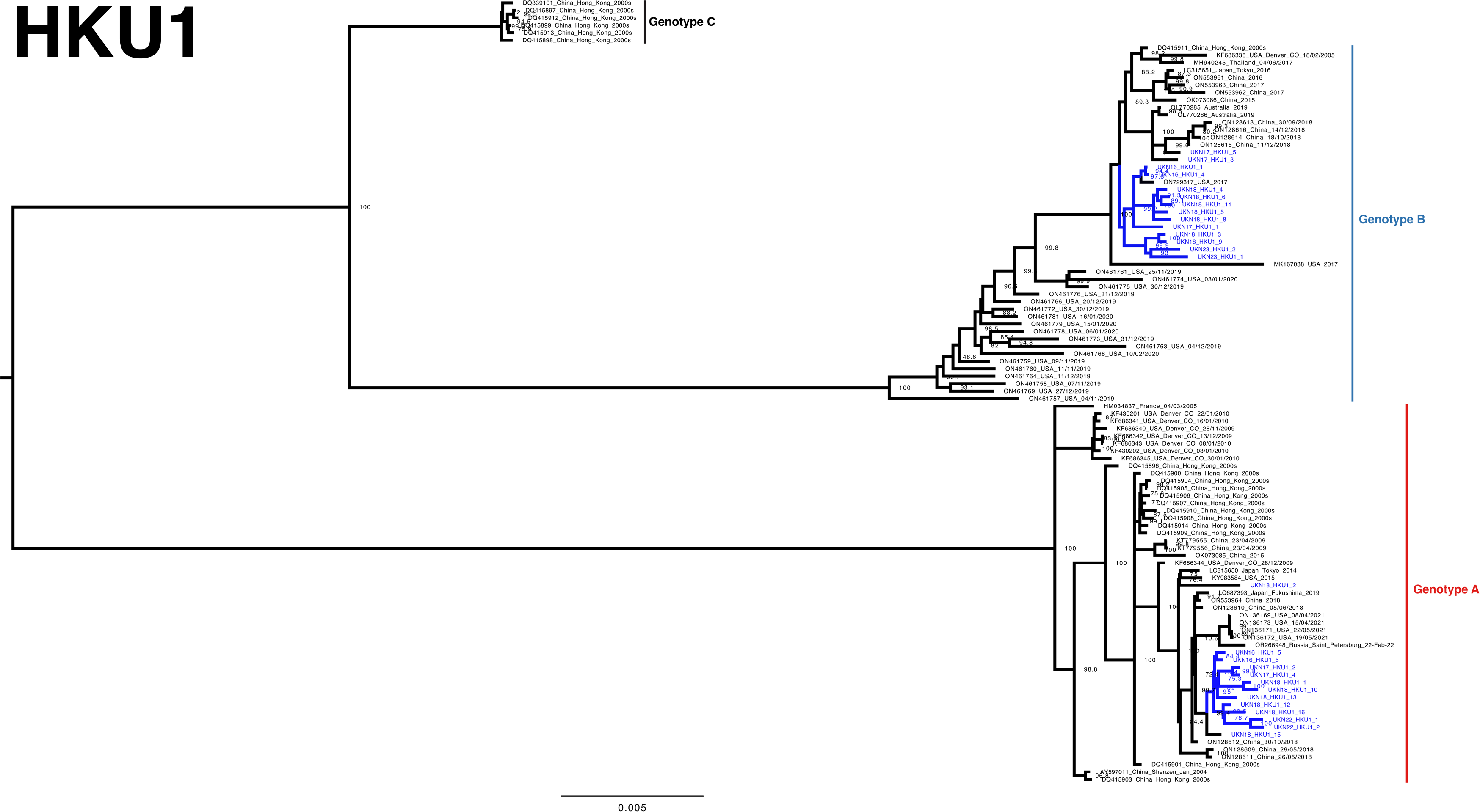
Phylogeny of HKU1 Seasonal Coronavirus. Phylogenetic relationship by maximum-likelihood method of all Nottingham, UK, study se-quences with >95% coverage (blue text) and publicly available genomes (black text) retrieved from GenBank in December 2023 (identified by acces-sion number/country [with region where noted]/year). Numbers to the right of tree nodes indicate SH-aLRT (Shimodaira–Hasegawa approximate like-lihood ratio test) branch support, with values <70 not shown. Branch lengths are drawn to a scale of nucleotide substitutions per site, with scale indi-cated. Tentative genotypes based on well supported clustering (>90 branch support) with lineage exemplar sequences as presented by Ye et al (16) are indicated by vertical bars.

**Figure 6:**
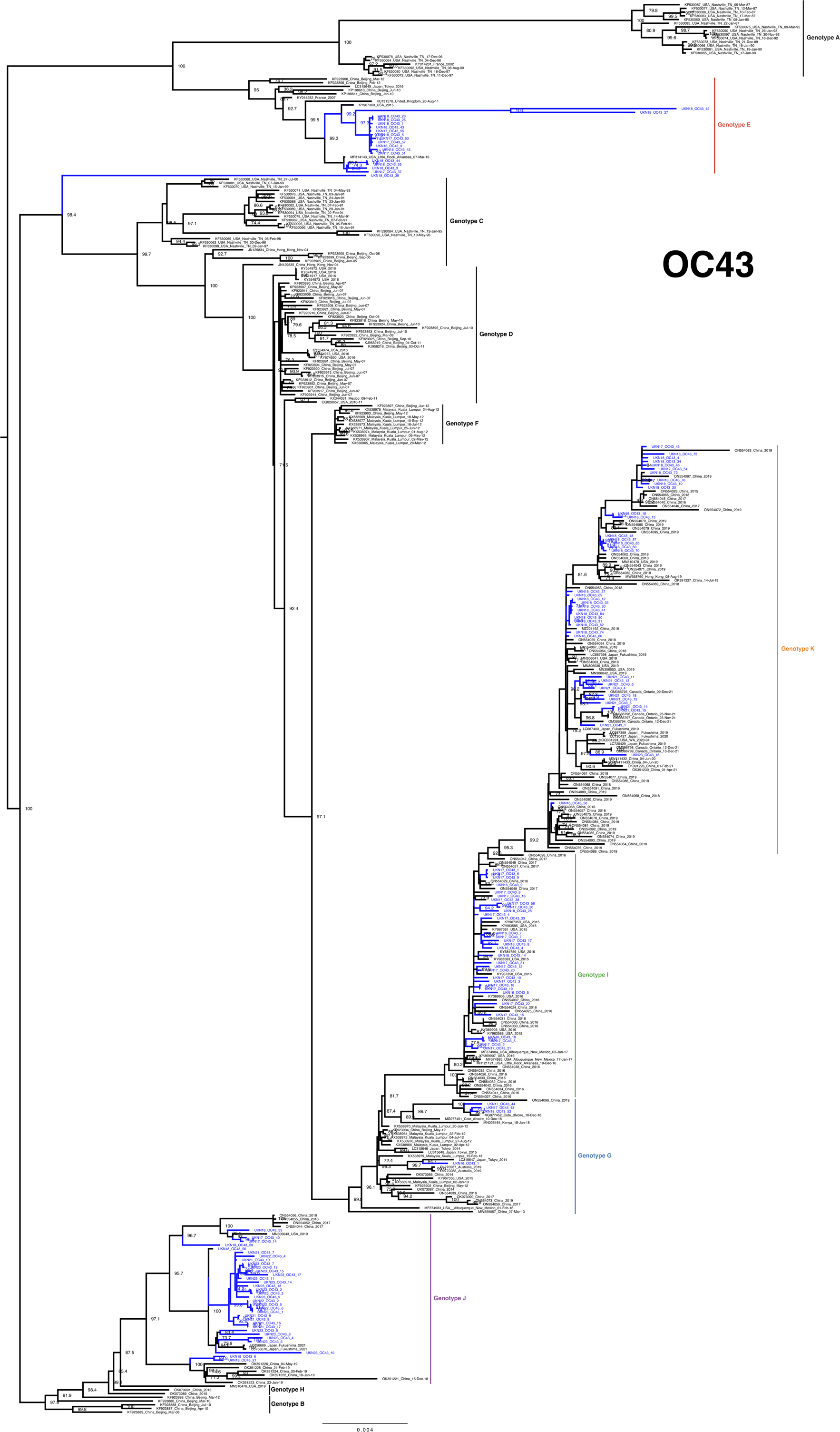
Phylogeny of OC43 Seasonal Coronavirus. Phylogenetic relationship by maximum-likelihood method of all Nottingham, UK, study sequences with >95% cov-erage (blue text) and publicly available genomes (black text) retrieved from GenBank in December 2023 (identified by accession number/country [with region where not-ed]/year). Numbers to the right of tree nodes indicate SH-aLRT (Shimodaira–Hasegawa approximate likelihood ratio test) branch support, with values <70 not shown. Branch lengths are drawn to a scale of nucleotide substitutions per site, with scale indicated. Tentative genotypes based on well supported clustering (>90 branch support) with lineage exemplar sequences as presented by Ye et al (16) are indicated by vertical bars.

### 229E

Alphacoronaviral 229E study sequences broadly group into 3 well supported clades (Figure 3, genotypes 6 / 7a / 7b). Three sequences from 2016 and one from January 2017 cluster with contemporary samples from the USA (2016 & 2017) and Germany (2015) and Vietnam (2013) assigned to Genotype 6 in previous studies (11, 16). Sparse sequence number representation, relatively long branch lengths and additional well-supported nodes is suggestive of an unsampled diversity within 229E genotype 6. However, the bulk of study sequences situate in a previously noted emerging lineage 7 (16) and are further segregated into two well supported and populated clusters, nominally 7a and 7b, both of which contain study genomes from 2016.

Interestingly, putative genotype 7a was comprised predominantly from our UK study sequences and none from the most extensive prior global study of 229E genomes, set in China immediately preceding the pandemic (16). Genotype 7a was only sampled elsewhere in the USA between 2015 and 2017, and not after the summer of 2018 in our study having been the significant majority variant in the core 2016/17 coronaviral season. In contrast, Genotype 7b was also seen from the outset of our study period, in samples derived from the USA and the key pre-pandemic Chinese Study (16), as well as post-SARS-CoV-2 emergence in Nottingham as well as China, Russia and the USA.

Increasing phylogenetic granularity, we can observe a single UK study sequence UKN18_229E_4 in June from a patient in their 20s, outside of typical UK coronaviral season) sitting within an extensive sub-cluster of Chinese sequences. Similarly, the earliest study sequence UKN16_229E_1 is more closely related to 2017 and 2018 Chinese sequences than anything else from the UK. Conversely, well supported and populated 7b sub-clusters of predominantly UK sequences with accordingly sparse Chinese representation can be observed. More recent 2022 and 2023 study sequences are found in these distinct genotype 7b clades.

Taken together, the genetic epidemiology of 229E supports the previously suggested notion of genetic drift (36), potentially with local lineage displacement of 7a by 7b suggesting a more nuanced segregation of geographical transmission at regional levels.

### NL63

With the significant NL63 cohort presented here and elsewhere recently (16), previously undetected diversity in circulating NL63 could also be observed. Diverse B and C lineages were sequenced throughout the study timeline (Figure 4), with many subtypes detected within the most heavily sampled 2017/18 core season (Figure 7). Notably in lineage B, the increased sampling supports an expansion of nomenclature toward designation of subtypes, similar to lineage C (37). We have tentatively annotated the well supported lineage B clusters subtypes B1 and B2 to assist with discrimination here and potentially elsewhere.

**Figure 7:**
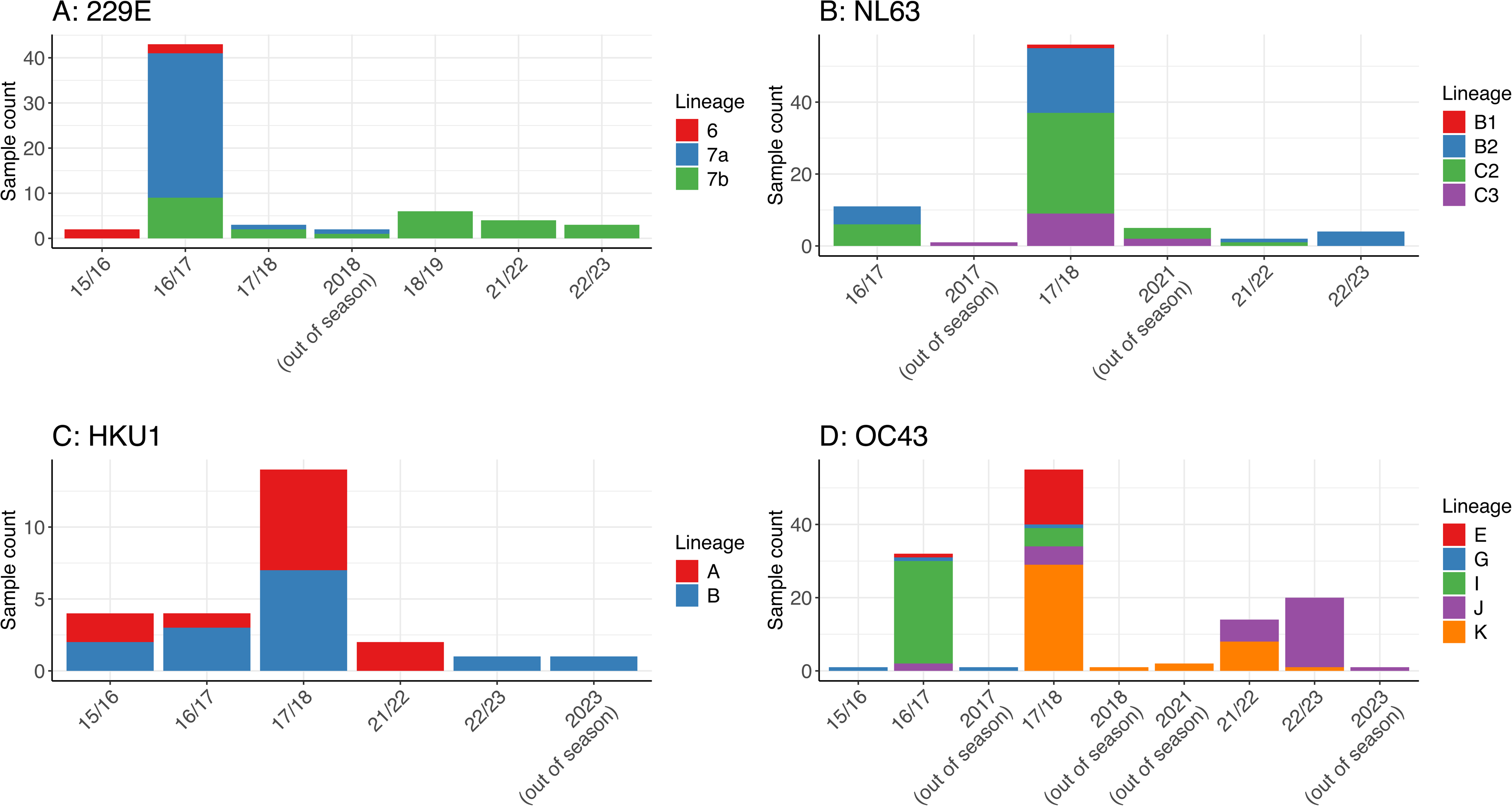
Lineages counts of Seasonal Coronaviruses by season. The lineages of 229E, NL63, HKU1 & OC43 Seasonal Coronaviruses (panels A to D respectively) were determined for each genome sequence of greater than 95% coverage, based on well supported clustering (>90 branch support) with lineage exemplar sequences as presented by Ye et al (16). Core infection season was defined as October to May (e.g. 16/17 for samples collected between October 2016 and May 2017), whilst June to September was considered out of typical season (e.g. 2017 for a sequence generated in June 2017).

Like its counterpart Alphacoronavirus 229E, NL63 exhibits geographical segregation within its subtypes. For instance UKN17_NL63_22, situating in a well-supported subgroup of B1 with reference sequences from China (2016–2019) and Japan (2018), whilst the bulk of B1, the ‘emerging cluster’ reported by Ye et al (16) with many 2016-2019 Chinese samples, contains no other UK study sequences despite our extensive sampling in a similar timeframe. Within Lineage B, our UK sampling presents most commonly as B2, with only single representatives from the USA (2015) and Japan (2017), in contrast to multiple 2017, 2018, 2022 and 2023 UK study sequences.

A similar picture presents with lineage C2, indicating a well-supported sub-cluster containing predominantly study sequences from 2016-18 and 2021/22, alongside contemporary US (2019) Japanese (2019) and Russian (2022) references. Elsewhere more limited numbers of further C2 and C3 sequences were observed pre- and post-pandemic, alongside more numerous Chinese and Japanese reference samples. Intriguingly a single study sample UKN18_NL63_24 stands distinctly from all others, Figure 4), potentially representing a novel intra-species recombinant of lineages B and C as extensively inferred previously (17, 38).

### HKU1

HKU1 is the least sequenced Seasonal Coronavirus globally due to an accordingly lower detection rate, therefore even our relatively limited contribution of 27 high quality (>95% coverage) genomes is significant (Figure 5). Genotypes A (n=13) and B (N=14) were detected in approximately equal numbers across the study period, presenting in sub-clusters with other contemporary sequences; we did not detect the recombinant, and to date rarest, Genotype C (14, 39).

Genotype A study samples from 2016-18 and 2022 segregated with high bootstrap support alongside contemporary Japanese (2014 & 2019), US (2015 & 2021) Chinese (2018) and Russian (2022), hinting at extinction of more historically detected genotype A lineages. Genotype B also offers only limited granularity currently, although the two most recent 2023 study samples segregate with earlier UK study genomes, separate from a cluster of US sequences seen at the onset of the SARS-CoV-2 pandemic (28). Other Genotype B study sequences appear more similar to contemporary Chinese (2015–18), Japanese (2016), Australian (2019), Thai (2017) and US (2017, with a temporally outlying 2005 sequence) isolates.

### OC43

OC43 is the most extensively sequenced seasonal coronavirus globally, contributing toward a more complicated genotypic nomenclature. We observed study sequences segregating with genotypes E, G, I, J and K both across the entire study and all within the most sampled 8-month core season of 2017/18 (Figure 6 and 7). The seasonal lineage data is again suggestive in the better-sampled pre-Pandemic era of a predominance in 2016/17 of genotype I being displaced in the following 2017/2018 season by the emergent genotype K (15). Post-Pandemic, a parity of genotype K with the additional emerging genotype J (15) in the 2021/22 season gives way to a predominance of genotype J in the most recent core season of 2022/2023. Interestingly, sample UKN18_OC43_38 stands alone on a long well supported branch, suggestive of further unsampled diversity in circulating viruses.

Intragenotypic temporal geographical segregation can again be observed, with Genotype E infrequently detected elsewhere globally, but featuring as the second most abundant lineage in our most extensively sampled 2017/18 season (Figure 6 and 7), in stark contrast to an absence of genotype E in 74 contemporary reference genomes from China (16). Genotype J indicated some additional well supported clades, potentially indicative of further emerging lineages set to predominate in future seasons.

### Prolonged infection by both Seasonal Alpha- and Betacoronaviruses

Several patients presented multiple samples of the same seasonal coronavirus type, although all but two of the serially samplings were within a 3-week timeframe, and most within a few days (data not shown). However, one patient (in their 20s) was determined to be NL63-positive over 195 days and 7 consecutive timepoints in 2017 (UKN17_NL63_1, 6, 7, 15, 16, 18 & 19), whilst another patient (in their 30s) was OC43-positive over 164 days and 4 consecutive timepoints in 2023 (UKN23_OC43_7, 12, 15 & 17). The sequential samples clustered closely on their respective trees (Figures 4 and 6) Both patients were noted to have been under respiratory surveillance post-bone-marrow transplant and thus highly likely to be immunosuppressed. Two weeks after the final positive timeline, the OC43-positive individual was negative for all targets in the viral respiratory multiplex RT-qPCR. The NL63-positive patient was not screened again after the final positive timepoint. The proximity of such genetically related positive samples suggested a prolonged infection as previously presented with SARS-CoV-2 (40), so the sequence differences were investigated in isolation for intrapatient evolution (Figure 8). The Alphacoronaviral NL63 patient presented an identical genome 22 days after the initial detection, however as the infection continued, 18 mutations arose in the ORF1ab (n = 3, 2 non-synonymous [NS]), Spike (8, all NS), NS3 (2, 1 NS), Matrix (1 NS), and Nucleocapsid (4, 2 NS and a 9bp). Half of the mutations, involving all genes and including the 3 amino acid nucleocapsid deletion appeared in the last time point.

**Figure 8:**
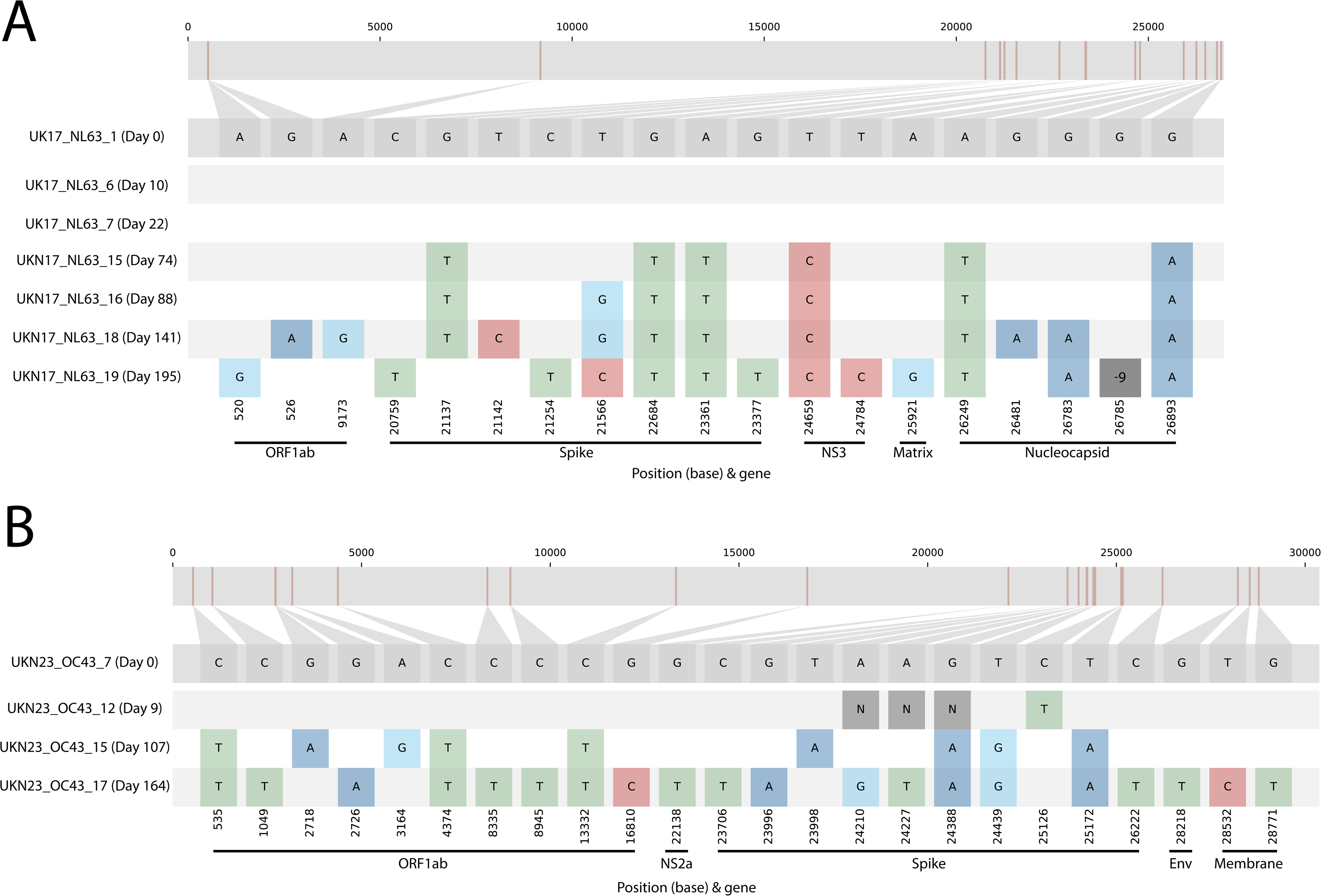
Putative prolonged Seasonal Coronaviral infection. Two sets of genome sequences were derived from the same individuals on dates suggestive of prolonged single infections, for both Alphacoronavirus NL63 (panel A) and Betacoronavirus OC43 (panel B). Sequence differences were visualised by Snip-it plots from the CIVET tool (https://github.com/artic-network/civet), using the first sample as the ‘Day 0’ timepoint and reference sequence. Genome nucleotide positions and genes are indicated relative to reference sequences LC488390 and LC756670 for NL63 and OC43 respectively. ‘N’ indicates sequence read dropout and ‘-9’ a 9bp deletion in the final NL63 nucleocapsid sequence.

Similar early and late time course mutation patterns were observed for the Betacoronavirus OC43 with no differences seen in the initial acute phase at 9 days. In contrast, mutations at 9 sites were observed at the 107-day timepoint and 14 further at 164 days, in the ORF1ab (n=10, 8 NS), NS2a (1 NS), Spike (10, 8 NS), Envelope (1 NS) and Membrane (2 NS).

### Co- and re-infected patients

In addition to the above mentioned OC43 / SARS-CoV-2 co-infected patient, the diagnostic testing indicated a further individual positive for both Alpha- and Betacoronaviruses (NL63 and HKU1), previously undetected by the non-discriminatory diagnostic assay, with a combined seasonal coronavirus genomic template value of 1771 copies / 10µl. A complete HKU1 genome and 47% of the NL63 genome was recovered by the Full Vivaldi method, elevated to 82% by subsequent NL63-exclusive sequencing.

Further to the consecutive timepoint NL63- and OC43-infections, 4 instances of multiple heterologous coronaviral infection in the same season were observed, all with NL63 and OC43 (data not shown). These ranged from as little as 38 to as many as 128 days apart, and all were observed within the most heavily sampled 2017/18 season.

## Discussion

The scale and rapidity of the sequencing efforts since the start of the SARS-CoV2 pandemic (41, 42), accompanied by the extensive media coverage and its impact on government health policies, has underscored the importance of viral genomics. While there has been significant global attention on this novel coronavirus spillover, there has been a notable absence of genomic data for the four endemic seasonal coronaviruses. This represents a critical gap in our knowledge that could provide insights into the future course of SARS-CoV-2 in the human population (35, 43, 44). Our study delivers a simple method to improve ongoing surveillance of seasonal coronaviruses and contributes substantially by providing ‘backfill’ of recent pre-pandemic genomes of all four species.

To reproducibly detect whole genomes from seasonal coronavirus strains (45–47) in unenriched clinical samples, demanded not only an amplicon approach, but also a fine balance between minimising priming sites and maximising the sensitivity of detecting smaller RNA fragments. We thus opted for a 1200bp amplicon scheme as used previously for SARS-CoV-2 (20), rather than the commonly adopted 400bp amplicon scheme (19, 21), larger amplicon schemes (18, 23, 24) or an unbiased metagenomic approach (28, 48). Initial attempts on even a small batch of clinical samples can quickly identify amplicon drop-out, whilst careful continued evaluation is required to guard against evolving drop-out such as seen previously in SARS-CoV-2 (25), and similarly seen in our cohort in post-pandemic 229E infections.

This approach has high sensitivity, with whole genome retrieval from samples with less than 100 genome copies of input, in line with previously reported sensitivity of the 400bp amplicon of 50 copies per reaction (21) and likely orders of magnitude more sensitive than metagenomic approaches to coronaviral genome sequencing (28). The comparable sensitivity of our larger 1200bp to previous 400bp schemes can be explained in part by increasing amplification cycle number from the typically used 35, to 45 (data not shown, (20, 21)). Concerns about the introduction of PCR error and / or detection of contaminating sequences by elevated cycle number can be minimised by not only consistent use of negative controls (49), but also use of proof-reading enzymes (19). The minimal differences observed in our closely but independently sampled OC43 infections – 4 differences across 6 paired c. 30Kb genomes – indicates a robust degree of fidelity in the method but this could be further tested with clonal templates to probe error rates. Similarly, to metagenomic sequencing, strong signals of only fractional parts of a coronaviral genome should be treated with suspicion (50).

To maximise the scope of our method, we ensured our scheme could target an unprecedented 5 near-complete species’ genomes from two coronaviral genera in the minimal 2 reaction format (21). To our knowledge, this is the broadest complete genome amplicon method presented to date and showcases the considerable potential of the method in sensitively targeting multiple viruses in whole, or part (51). We validated this by confirming clinically diagnosed coronavirus co-infections and identifying others missed by RT-qPCR multiplex assay panels, which can have poor discriminatory potential.

Combining our novel methodology and extensive diagnostic surplus archive, we have generated the largest single cohort of seasonal coronaviral genomes to date. Thus, when combined with the global dataset, and notably other contemporary in-depth studies from China (16) and the US (28), we were able to gain greater insight into the ebb and flow of seasonal coronaviral genotypes / lineages, such as has been monitored with unprecedented detail with the analogous SARS-CoV-2 variants. In contrast to the currently observed pattern of emergence and apparent total selective sweeps of SARS-CoV-2 lineages (35, 52)(REF), the seasonal coronaviruses were observed to present a more complex co-circulation of genetically distinct lineages rising and falling in predominance, sometimes with contemporary geographical variation and prevalence. For example, 229E saw a progressive sweep in our UK cohort through lineages 6 and 7a pre-pandemic, to exclusively 7b post-pandemic; however 7b was predominant in a pre-pandemic Chinese cohort (16). Similarly, OC43 lineage I was displaced in predominance by the closely genetically related lineage K (15) between the 2016/17 and 2017/18 seasons. Whilst lineage J, a variant of precursor lineage H (15) was also present in a minority in both these seasons, it presented as the overwhelming OC43 lineage of the 2022/23 UK season. It was generally notable that the season we sampled most extensively (2017/2018) also exhibited the greatest diversity of both alphacoronaviral NL63 and betacoronaviral OC43 lineages, further emphasising the need for greater genomic surveillance to capture the true extent of circulating seasonal coronavirus variability. Underlining this point was the high prevalence of OC43 genotype E in 2017/18, infrequently observed elsewhere globally in recent sampling, but strikingly associated with a fatal encephalitis case previously also in the UK in 2011 (53).

It has been noted for NL63 that genotype switching was not a prerequisite to reinfection of an individual (48), supported by phylogenetic analysis and in contrast to 229E and OC43 (44). The selective force on 229E to antigenically evolve has been further demonstrated *in vivo* with historical sera failing to neutralise more contemporary isolates through significant variation in receptor-binding domains (43) with our data indicating the trend continues. Although an older study suggested re-infection by the same 229E strain was indeed possible in the short term (54), this was in a highly controlled experimental setting and in direct contradiction to a similar earlier study in the 1980s (55). As noted elsewhere (44), HKU1 data is insufficient to draw conclusions about its evolutionary trajectory, however the novel method and relatively large HKU1 cohorts presented here and recently elsewhere (28) could redress this imbalance in future studies. However more broadly, absence of a recombinant HKU1 A / B lineage C extant around the time HKU1 was discovered (6, 14) was in agreement with other key contemporary studies (28, 39).

Overall, this may suggest that dominating SARS-CoV-2 lineages in well-sampled region may not subsequently be swept to extinction but could continue undetected circulation and further evolution in certain populations before re-emergence in others. The unexpected appearance and subsequent domination of novel SARS-CoV-2 variants Delta and Omicron were indeed suggestive of transmission and evolution in unsampled populations (42, 56, 57) or longer term intra-host evolution in immunosuppressed individuals as discussed below. The significant differences between the genomes in this single centre study and the other largest contemporary seasonal coronavirus cohort to date, from 36 hospitals in Beijing with a catchment of 25 million people (16), underscores the need for greater and broader surveillances of respiratory viral infections, not only for seasonal coronaviruses, but other pathogens also (58).

The effect of non-pharmaceutical interventions to reduce pandemic transmission of SARS-CoV-2 has had a significant effect on the well-monitored circulation of Influenza, notably with the apparent disappearance of Influenza B Yamagata (59). These measures may also have exerted pressure on some seasonal coronavirus lineages, with a significant delay in the typical US seasonal onset observed in 2020/21 attributed to the SARS-CoV-2 pandemic (10). Pre-pandemic patterns of alternating seasonal betacoronaviral dominance of OC43 and HKU1, punctuated by parity (10), may be further perturbed by significantly enhanced population immunity (by infection and / or vaccination) of SARS-CoV-2 as a third beta-coronavirus (60). But ultimately immunity against re-infection, cross-protective or otherwise, is short-lived (61), whilst protection against severe disease remains (60, 62).

Paradoxically ‘out of season’ seasonal coronaviral infections were observed. In some instances, phylogenetic analysis was suggestive of importation to the UK by international travel, as we and others reported previously for SARS-CoV-2 (41, 42). Conversely, there may be continued low-level circulation within the UK and elsewhere. In other instances, out of season positives were the result of likely persistent infections acquired by the immunocompromised within the typical epidemic months. Long-term SARS-CoV2 infection of individuals (especially immune-suppressed), leading to intra-patient virus evolution has been described (40, 63), and offered as a potential mechanism for the emergence of some highly evolved variants of concern (63, 64). We similarly demonstrate that increased genomic sequencing of seasonal coronaviruses in even our single centre cohort can reveal prolonged infection and evolution in both beta- and alphacoronaviruses. Perhaps most strikingly we observed a 9bp nucleotide deletion immediately prior to the start of the C-terminal domain of the nucleocapsid in the disordered linker region responsible for RNA binding and oligomerisation (65, 66) in the last timepoint of our chronically infected NL63 patient. This had parallels in SARS-CoV-2, with a 3 AA nucleocapsid deletion relative to the ancestral Wuhan strain a feature of the Omicron lineage, albeit in the N terminal domain (67). The recent spillover of a novel canine coronavirus in Malaysia also reported a larger 12 AA deletion in the middle of the N protein, hypothesised to be associated with adaptation to a new host (68), similar to previously reported nucleocapsid deletions (and insertions) found to determine nuclear localization in MERS and SARS-CoV-1 infections (69). However, caution must be taken in extrapolating any isolated changes in immunocompromised patients to effects on transmissibility in the wider population.

In conclusion, given the recent spillover of SARS-CoV-2, its uncertain evolutionary path, the concurrent circulation and potential co-infection with endemic coronaviruses, and the propensity of coronaviruses for genetic recombination, there is a pressing need for enhanced surveillance of seasonal coronaviruses. The methodology outlined in this study closely resembles widely accepted amplicon schemes used for SARS-CoV-2, but with a notable advancement: it offers unprecedented coverage across two genera and five species. Furthermore, it has demonstrated high sensitivity in both pre- and post-pandemic cohorts of all four seasonal coronaviruses. Therefore, it is well-suited for widespread adoption, serving as a valuable tool to complement the increasing genomic sequencing efforts of other respiratory viruses. This will provide crucial insights for informed epidemiological analyses and public health decision-making.

## Ethical Approval statement

The COG-UK consortium work was approved by the PHE Research Ethics and Governance Group (R&D NR0195). Use of pre-pandemic residual diagnostic nucleic acids and associated anonymized patient information was covered by ethical approval granted to the Nottingham Health Science Biobank Research Tissue Bank, by the North West -Greater Manchester Central Research Ethics Committee, UK, reference 15/NW/0685. Nottingham University Hospitals National Health Service Trust (Nottingham, UK) further approved investigation of diagnosed coronavirus positive samples under clinical audit number 23-078C.

## Data Availability

All data produced in the present study are available upon reasonable request to the authors

---

**Suppl. Figure 1:**
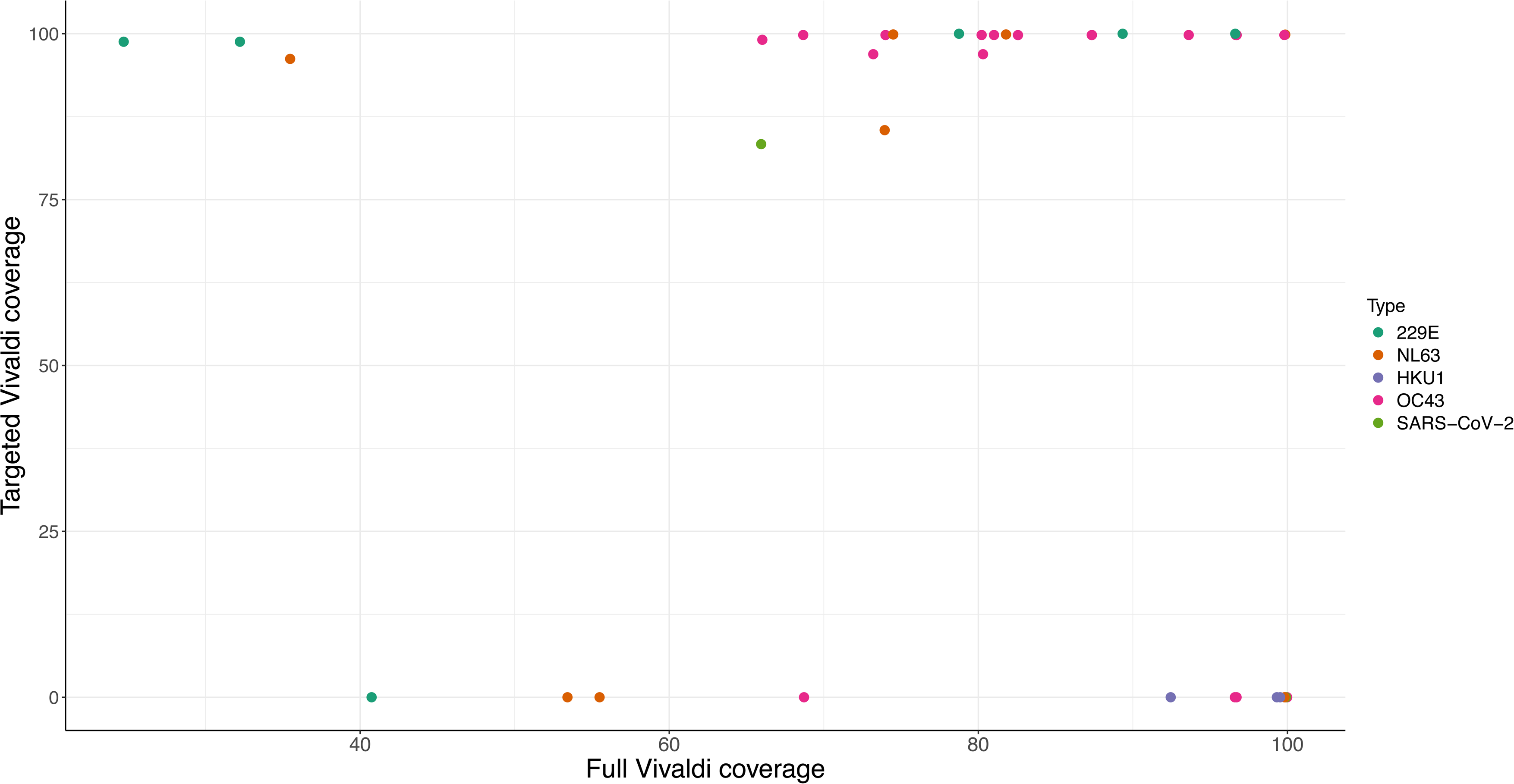
Genome coverage comparing Full Vivaldi four seasonal coronaviruses and SARS-CoV-2 combined multiplex to type-matched, targeted sequencing. The genome coverage of selected Seasonal Coronavirus 229E, NL63, HKU1 & OC43 and additionally SARS-CoV-2 samples when amplified by PCR multiplex containing all primer sets (Full Vivaldi, x axis), is shown compared to the genome coverage achieved with a single set of coronavirus-targeting primers (Targeted Vivaldi, y-axis). Genome coverage is expressed as a percentage of the total region targeted by the amplicon scheme. Samples with ‘0’ coverage by the targeted method were sequenced only by the Full Vivaldi method.

